# Adverse Events in Surgical Neurology: The Novel Therapy-Disability-Neurology (TDN) Grade

**DOI:** 10.1101/2020.07.06.20144824

**Authors:** Alexis Paul Romain Terrapon, Costanza Maria Zattra, Stefanos Voglis, Julia Velz, Flavio Vasella, Kevin Akeret, Ulrike Held, Silvia Schiavolin, Oliver Bozinov, Paolo Ferroli, Morgan Broggi, Johannes Sarnthein, Luca Regli, Marian Christoph Neidert

## Abstract

**Background:** The most widely used classifications of adverse events (AE) in surgical neurology assign a grade to AE that depends on the therapy used to treat them or on new neurologic deficits. Both concepts have substantial shortcomings in grading AE severity. We present a novel multidimensional approach to this challenge and aim at validating the new grading system.

**Methods:** The new Therapy-Disability-Neurology (TDN) grading system classifies AE into five grades, depending on the associated therapy, disability, and neurologic deficits. We conducted a two-center study on 6071 interventions covering the whole neurosurgical spectrum with data prospectively recorded between January 2013 and September 2019 at the University Hospital Zurich (USZ) and at the Fondazione IRCCS Istituto Neurologico Carlo Besta (FINCB).

**Findings:** Using data from USZ, a positive correlation was found between the severity of AE and the length of hospital stay (LOS) as well as treatment cost. Each grade was associated with a greater deterioration of the Karnofsky Performance Status Scale (KPS) at discharge and at follow-up. Additionally, there was a correlation between the severity of AE and absolute KPS values. When using the same methods on an external validation cohort from FINCB, correlations between the grade of AE, LOS, and KPS at discharge were even more pronounced.

**Interpretation:** Our results suggest that the TDN grade is consistent with clinical and economic repercussions of AE and thus reflects AE severity. It is objective, practical, easily interpreted, and enables comparison between different medical centers. The TDN grade will constitute an important step forward towards a more precise and standardized documentation of AE and ultimately lead to a more critical and patient-centered appraisal of process and outcome measures in surgical neurology.

**Funding:** None.

## INTRODUCTION

Technical innovation in surgical neurology is speedy and new methods are constantly being developed to guarantee patient safety, to minimize the occurrence of adverse events (AE), and ultimately to improve patient survival and quality of life. Unfortunately, an essential tool is still lacking: a widely accepted classification of the severity of AE specific to surgical neurology and carefully tailored to handle the functional complexity of the central nervous system. Such a tool would give the possibility to monitor, to compare, and to improve quality of the treatment delivered. In 2011, Clark and Spetzler defined the characteristics of an ideal classification scheme:^1^ it should enable to recognize and adequately quantify the severity of AE while staying functional, unbiased and easily interpreted. They suggested that such an invention would be a critical step in the creation of a “brave new world for neurosurgery”.

Different classifications have been developed, but none has gained wide acceptance. The most reported is the therapy-based five-grade Clavien-Dindo-Grading system (CDG) proposed for general surgery in 2004.^2-5^ Landriel et al.^6^ later adapted this system to cranial and spinal neurosurgery, but the Landriel Ibañez Classification (LIC) remains very similar to CDG and one can easily be translated into the other. Despite being excellent in quantifying which resources are needed to treat an AE, these methods contain limitations which are particularly important in our field, as they fail to detect the severity of AE that are not treated by means of pharmacotherapy and/or surgery. A new neurologic deficit, for instance, is a frequent AE occurring after neurosurgical procedures.^6-11^ Although they may imply dramatic consequences on quality of life,^12^ such events are usually considered as low grade in grading schemes that are solely therapy-based; such concerns have repeatedly been expressed in the literature.^2,8,13-16^ Scales that assess disability such as the modified Rankin Scale (mRS),^17^ or neurologic deficits such as the National Institutes of Health Stroke Scale (NIHSS) have been shown to be good predictors of the quality of life months after central nervous system lesions.^18^ We believe that these factors are of great clinical and prognostic importance and should be considered, along with the therapeutic resources involved, to fully capture the severity of AE. We therefore propose a multidimensional and patient-centered classification scheme. By investigating the relationship between the grade of AE and the duration, cost, and clinical outcome of neurosurgical management in a cohort from our prospective patient registry,^2^ we first assess the applicability of the new grading system. Next, we validate its applicability on an independent cohort from a different country.^13,19^ We describe, challenge, and discuss our scheme with the aim to provide the ideal classification of the severity of AE in surgical neurology.

## METHODS

### Patients

All patients that underwent elective and urgent neurosurgical procedures at the University Hospital Zurich (USZ) between January 2013 and August 2019 with the completion of data entry (including the identification number of the operation, the indication for surgery, the date of operation, admission, discharge, and birth) in a prospective patient registry on admission and upon discharge were considered. From this cohort, we removed all interventions for which the mRS and the Karnofsky Performance Status Scale (KPS) were not known at the time of admission or discharge. If the same patient underwent more than one surgical procedure within three months, only the first was included. For the section of our study where the impact of AE at follow-up was explored, we further excluded all interventions that were not tracked with mRS and KPS within three months. Following the same criteria, the validation cohort was selected from patients that underwent elective neurosurgical procedures at the Fondazione IRCCS Istituto Neurologico Carlo Besta of Milan, Italy (FINCB) between October 2017 and September 2019.

### Measures

Data were prospectively recorded by neurosurgeons at two independent neurosurgical centers and entered in separate registries that have been described earlier.^2,13,19,20^ In both centers, all data were retrospectively reviewed by board certified neurosurgeons. At USZ, every new team member was provided with introductory teaching and standardized written instructions and required to obtain a certificate for the correct use of clinical scores. In addition, all AE were validated at the monthly department meeting and at the monthly morbidity and mortality meeting. Health status was evaluated with KPS and mRS. Length of stay (LOS) was measured from the day of surgery to the day of discharge. AE were defined and classified according to the CDG at USZ and to the LIC at FINCB and were accompanied by a short description. To simplify our investigations, we only report the most severe event at each timepoint. The data regarding follow-up at USZ refers to the last consultation that took place between six weeks and three months after the operation, including AE that may have occurred until then. At FINCB, data at follow-up was recorded only for patients who have had a craniotomy, and AE were recorded when occurring within 30 days of discharge. Costs were only recorded at USZ. To facilitate the interpretation of our data and to ensure comparability to other institutions and countries, we have divided the total patient-related cost of each case by the mean patient-related cost of all cases (relative cost).

### Novel classification of the severity of adverse events

AE were defined as any deviation from the normal postoperative course and were ordered in relation to the therapy, disability, and neurological deficits they involved (Table 1). Death of any reason occurring within 30 days following surgery was considered an AE. According to the new Therapy-Disability-Neurology (TDN) classification, a grade was attributed to each event using custom scripts in MATLAB with the algorithm illustrated in Figure 1. The therapeutic dimension was measured according to the worst CDG at USZ and LIC at FINCB at each point in time and the disability with the difference in mRS between admission and discharge, or admission and follow-up when the AE had occurred after discharge. New neurologic deficits were identified by searching for key words in the description of AE at USZ and prospectively identified at FINCB.

**Table 1.** The Therapy-Disability-Neurology grade of adverse events in surgical neurology. In the Therapy-Disability-Neurology (TDN) grade of adverse events in surgical neurology, three dimensions are evaluated. The therapy, based on the Clavien-Dindo-Grading system (CDG) or the Landriel Ibañez Classification (LIC),^3-6^ the disability, using the modified Rankin Scale (mRS),^17^ and the neurology, using a binary definition. When dimensions of therapy, disability and neurology are of different severity, the highest resulting grade should always be chosen. * The dimensions of disability and neurology should only be considered when these have deteriorated since the preoperative state. This impairment must result from the adverse event.

| TDN | Definition | Therapy | Disability* | Neurology* |
| --- | --- | --- | --- | --- |
| **Grade 1** | Any adverse event without the need for a treatment or an intervention, that does not impact daily life activities, and does not result in any new neurologic deficit. Allowed therapeutic modalities are drugs as antiemetics, antipyretics, analgetics, diuretics, electrolytes, physiotherapy and bedside opening of wound infections. | CDG I \| LIC Ia | mRS <2 | No new neurological deficit |
| **Grade 2** | Any adverse event requiring pharmacological treatment (including blood transfusions and total parenteral nutrition) or hindering at least one activity of daily living, or resulting in a new neurologic deficit. | CDG II \| LIC Ib | mRS 2-3 | Any new neurological deficit |
| **Grade 3** | Any adverse event requiring an invasive procedure or hindering walking, or preventing the patient from attending his own bodily needs. | CDG III \| LIC II | mRS 4 |  |
| **Grade 4** | Any life- threatening adverse event requiring a management in intensive care or leaving the patient bedridden, in need of constant help, incontinent. | CDG IV \| LIC III | mRS 5 |  |
| **Grade 5** | Any adverse event resulting in the death of a patient. | CDG V \| LIC IV | mRS 6 |  |

**Figure 1.**
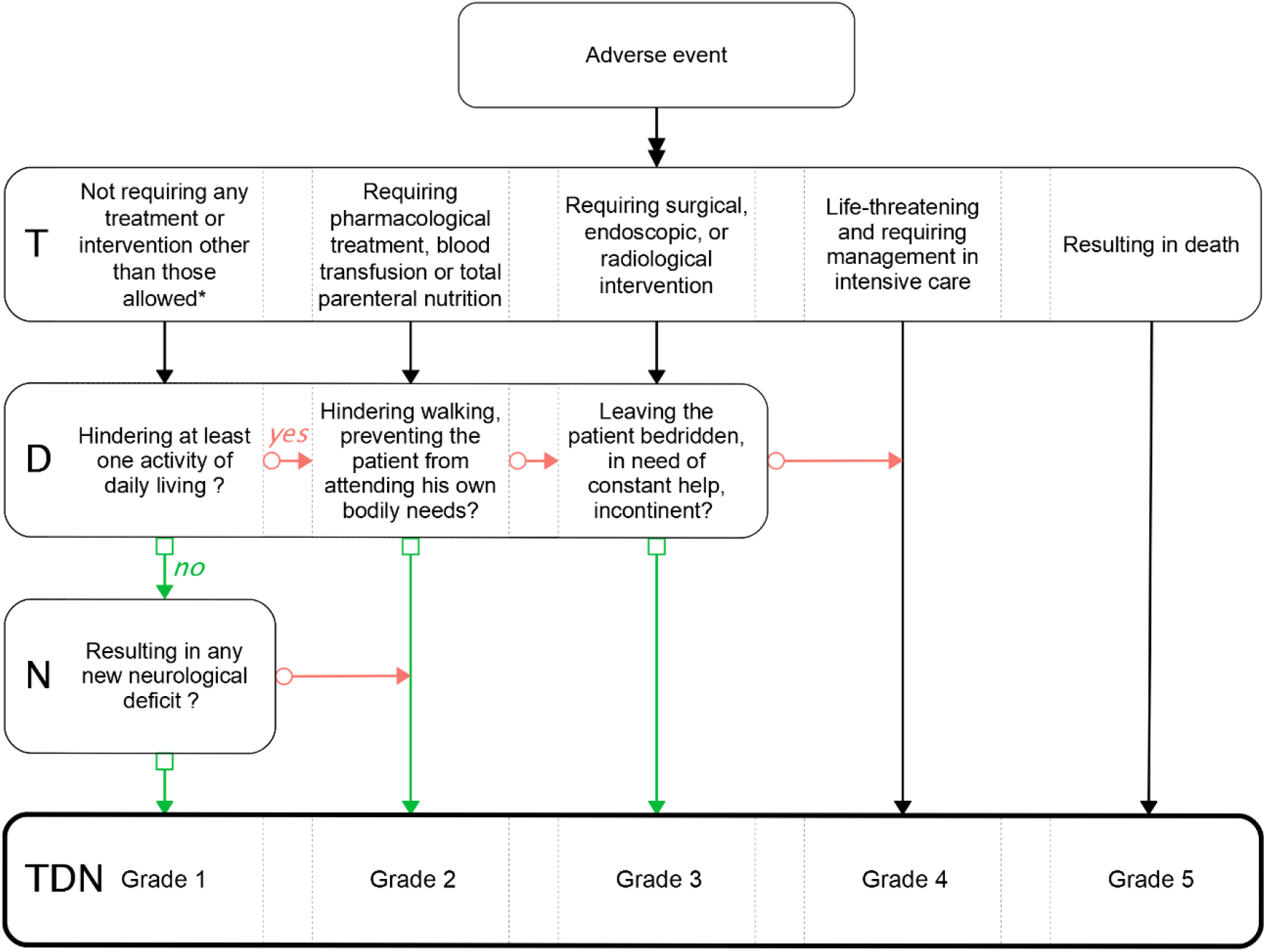
The Therapy-Disability-Neurology grade of adverse events in surgical neurology. In the Therapy-Disability-Neurology (TDN) grade of adverse events in surgical neurology, three dimensions are evaluated: Therapy (T), based on the Clavien-Dindo-Grading system (CDG) or the Landriel Ibañez Classification (LIC);^3-6^ disability (D), using the modified Rankin Scale (mRS);^17^ and neurology (N), using a binary definition. When the box of the flowchart contains a question, follow the red arrow to the right if the answer is “yes” and the green arrow down if the answer is “no”. For example, a patient with a new untreatable and non-disabling facial nerve paralysis will have a TDN grade of two (starting at the first column of T, the answer to the question of D is “no”, and the answer to the question of N is “yes”. *Allowed therapeutic modalities are antiemetics, antipyretics, analgesics, diuretics, electrolytes, and bedside opening of wound infections. Any other pharmacological therapeutic regimen are second grade severity criteria.

### Statistical analysis

We performed our statistical analysis and created figures using the statistical programming language R, including specific packages (see Supplementary Material). The cohorts from USZ and FINCB were kept separate in our analyses. We present means accompanied by standard deviations (SD), medians by interquartile range (IQR), and percentages by 95% confidence intervals (CI, Wilson method for binomial proportions and Sison-Glaz method for multinomial proportions). We compared variables between patients with and without AE using the t-test (age) or the Wilcoxon rank sum test (KPS, LOS, relative cost; reported with the median difference and the 95% confidence interval). We explored the difference in LOS, relative cost, and variation of KPS between multiple groups (AE grades) with the Kruskal-Wallis rank sum test; Dunn’s post hoc multiple comparison was used to compare the mean rank of each group with each other. Correlations were measured with Spearman’s rank correlation coefficient rho (reported with the proportion of shared variance R_s_^2^), except for the correlation between KPS and TDN grade, which was measured with Kendall’s tau. Multiple regression models (reported with the relative importance of each predictors normalized to sum to 100%) with each dimension of the TDN grade as separate predictors for LOS, relative cost, and KPS at discharge were compared to linear regression models using only the TDN as a predictor. The level of statistical significance was set to be < 0·05 and all reported p-values were corrected with the Benjamini-Hochberg procedure.

### Ethical considerations

USZ registry was approved upfront by the local ethics review board (PB-2017-00093) and internationally registered at clinicaltrials.gov (NCT01628406). Data recorded a FINCB was anonymized before our analysis. This study is reported in accordance with the Strengthening the Reporting of Observational Studies in Epidemiology (STROBE) statement.^21^

## RESULTS

We included and analyzed 4680 interventions performed at USZ between 2013 and 2019. We validated our findings in an independent cohort of 1391 interventions performed at FINCB between 2017 and 2019. Details regarding the number of patients at each stage of the study can be found in Supplementary Figure 1. At USZ, the mean age at surgery was 58 years (SD 17·71) and did not differ significantly between patients with AE (59 [18·32]) and without AE (58 [17·53], p = 0·0522) at discharge. We found evidence for a difference in KPS at admission between patients who would (median = 70% [IQR 50]) or would not (median = 80% [20]) suffer from an AE (median difference = 10% [10 - 10], p < 0·0001).

At discharge, we recorded 1007/4680 AE at USZ (21% [CI 20·36 - 22·71]) and 377/1391 at FINCB (27% [24·83 - 29·50]). Among all cases with AE at USZ, the proportion with low KPS as well as high LOS and costs rose with each TDN grade (Figure 2). Detailed results with the distributions of AE according to TDN, CDG and LIC are reported in Table 2. At USZ, the group of indication with the highest percentage of AE was vascular (259/796 cases, 33% [29·37 - 35·87]), followed by cerebrospinal fluid (115/387, 30% [25·38 - 34·45]), and trauma (146/572, 26% [22·12 - 29·25]); other results are listed in Supplementary Table 1. The distribution of TDN dimensions differed between each AE grade at USZ (Figure 3).

**Table 2.** Distribution of cases and median Karnofsky Performance Status Scale at admission and at discharge according to different classifications of adverse events. It should be noted that the University Hospital Zurich (USZ) cohort includes both elective and urgent operations, while the Fondazione IRCCS Istituto Neurologico Carlo Besta (FINCB) cohort includes elective operations only. Death of any reason occurring within 30 days following surgery was considered an AE. The median admission Karnofsky Performance Status Scale (KPS) is lower at USZ than at FINCB (elective only), and the median discharge KPS decreases with the increase in the Therapy-Disability-Neurology (TDN) grade in both institutions. Interquartile range (IQR), number (n). *The Clavien-Dindo-Grading system (CDG) and Landriel Ibañez Classification (LIC) have been merged to simplify reading.

|  | USZ |  |  | FINCB |  |  |
| --- | --- | --- | --- | --- | --- | --- |
| Grade | All cases | KPS median (IQR) |  | All cases | KPS median (IQR) |  |
| Adverse event | n (%) | Admission [%] | Discharge [%] | n (%) | Admission [%] | Discharge [%] |
| No | 3673 (78%) | 80 (20) | 90 (10) | 1014 (73%) | 90 (10) | 90 (10) |
| Yes | 1007 (22%) | 70 (50) | 70 (50) | 377 (27%) | 90 (10) | 80 (20) |
| **TDN** |  |  |  |  |  |  |
| 1 | 83 (8.2%) | 80 (20) | 90 (20) | 66 (18%) | 90 (10) | 90 (10) |
| 2 | 554 (55%) | 80 (40) | 80 (30) | 169 (45%) | 90 (10) | 80 (20) |
| 3 | 236 (23%) | 80 (40) | 50 (50) | 105 (28%) | 80 (20) | 70 (40) |
| 4 | 63 (6.3%) | 60 (40) | 40 (30) | 29 (7.7%) | 80 (20) | 50 (40) |
| 5 | 71 (7.1%) | 40 (45) | 0 (0) | 8 (2.1%) | 70 (20) | 0 (0) |
| **CDG (LIC)\*** |  |  |  |  |  |  |
| I (Ia) | 258 (26%) | 80 (20) | 80 (20) | 176 (47%) | 90 (10) | 80 (20) |
| II (Ib) | 446 (44%) | 70 (40) | 70 (40) | 82 (22%) | 90 (10) | 90 (20) |
| IIIa (IIa) | 46 (4.6%) | 70 (40) | 70 (48) | 31 (8.2%) | 80 (15) | 90 (15) |
| IIIb (IIb) | 147 (15%) | 70 (40) | 60 (50) | 57 (15%) | 80 (20) | 70 (30) |
| IVa (IIIa) | 36 (3.6%) | 65 (40) | 50 (33) | 18 (4.8%) | 80 (20) | 55 (40) |
| IVb (IIIb) | 3 (0.3%) | 90 (25) | 60 (30) | 5 (1.3%) | 70 (10) | 50 (10) |

**Figure 2.**
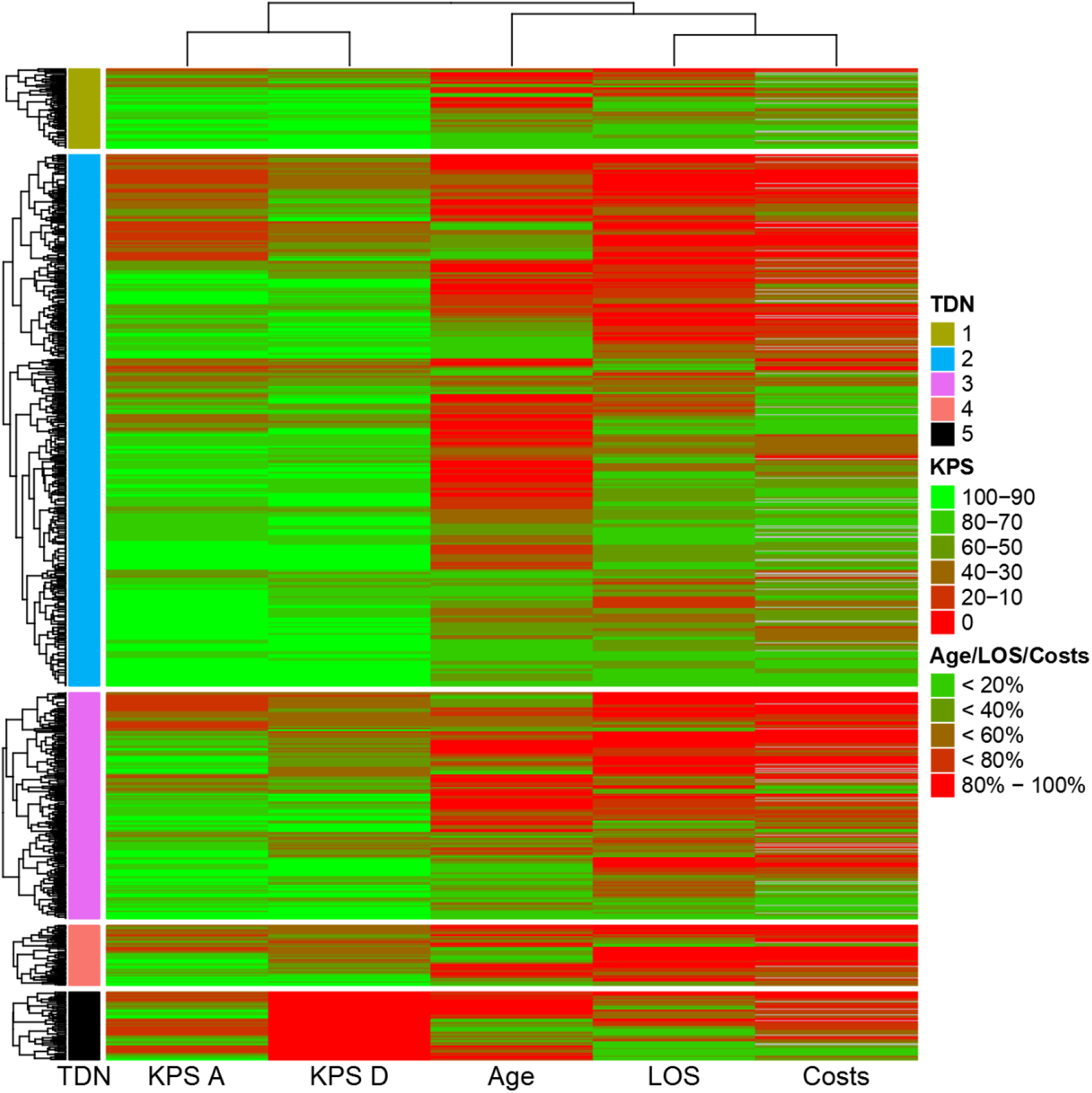
Heatmap with dendrogram split by Therapy-Disability-Neurology grade for cases operated at the University Hospital Zurich. All cases with an adverse event prior to discharge were included (1007 cases). In 85 cases, the associated costs were not known (grey). KPS at admission (KPS A) and KPS at discharge (KPS D) where divided into increments of 20. Age, length of stay (LOS), and costs were divided into quintiles. The heatmap was split according to the Therapy-Disability-Neurology (TDN) grade and the distance was measured using the Euclidean method. The proportion of cases with low KPS, high LOS, and high costs (red) rises with each TDN grade.

**Figure 3.**
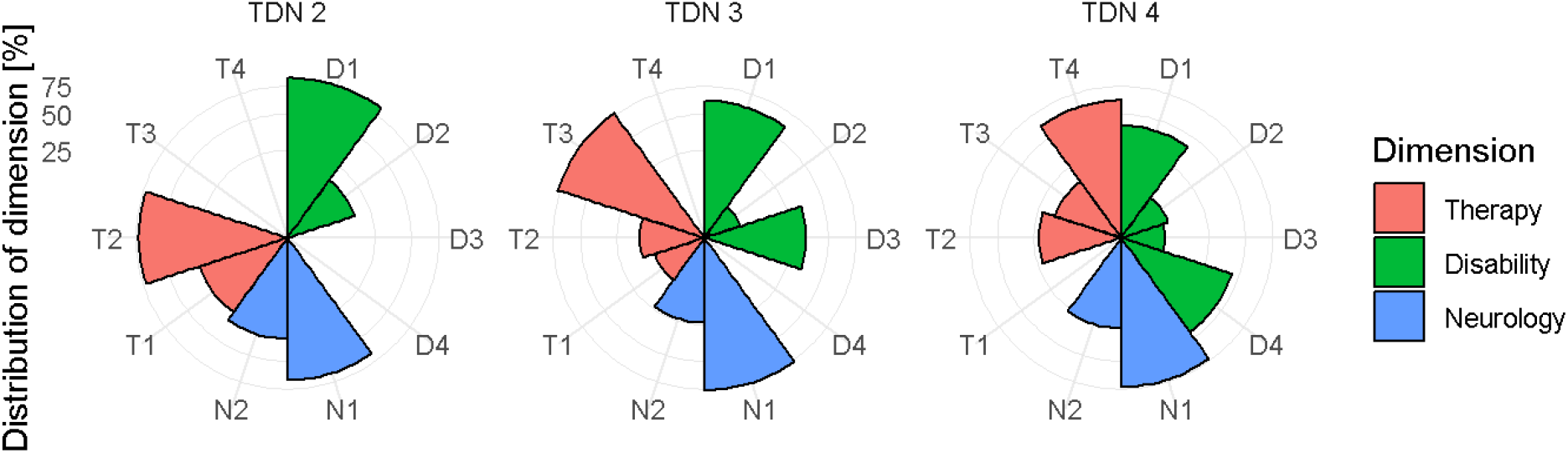
Distribution in percentage of the dimensions of the Therapy-Disability-Neurology grade at University Hospital Zurich. The letters followed by numbers indicate the grade of the adverse events based solely on the dimension being explored. For example, among Therapy-Disability-Neurology (TDN) grade 2 adverse events, 72% have a “therapy” (T) dimension of two (T2, requiring pharmacological treatment), 16% have a “disability” (D) dimension of two (D2, hindering at least one activity of daily living), and 34% have a “neurology” (N) dimension of two (N2, resulting in a new neurologic deficit). Note that the final grade is always derived from the dimension of greatest severity. Grade 1 adverse events always corresponds to T1, D1, and N1 (data not shown) and the dimensions cannot be addressed separately for grade 5 adverse events (death, data not shown). Only adverse events that occurred prior to discharge were considered.

At USZ, mean LOS was 7·7 days (SD 7·43) and median 6·0 (IQR 4·00). Mean relative cost was 1·0 (1·15) and median 0·67 (0·58). Hospitalizations associated with AE at discharge were much longer (mean = 14 days [11·61], median = 10 [11]) and more expensive (mean = 1·9 [1·86], median = 1·2 [1·59]) than those without AE (LOS: mean = 6·0 days [4·54], median = 5 [3], median difference = 5 [5 - 5], p < 0·0001; relative costs: mean = 0·72 [0·59], median = 0·59 [0·43], median difference = 0·58 [0·53 - 0·63], p < 0·0001). Both variables were influenced by the TDN grade of AE (p < 0·0001, Figure 4). A post hoc comparison between the mean ranks of each TDN grade and the event free group revealed that LOS and relative costs were gradually and significantly increasing with the severity of AE, except for grade 5 (death) as expected; detailed results are presented in Supplementary Table 2. There was a moderate positive correlation between TDN grade and LOS (rho = 0·42, R_s_^2^ = 17%, p < 0·0001) and between TDN grade and relative cost (rho = 0·45, R_s_^2^ = 20%, p < 0·0001). To limit the influence of the correlation between relative cost and LOS on our results (rho = 0·65, R_s_^2^ = 43%, p < 0·0001), we performed the partial correlation between TDN grade and relative cost by controlling for LOS and witnessed a decrease of Spearman’s rho (rho = 0·25, R_s_^2^ = 6.0%, p < 0·0001).

**Figure 4.**
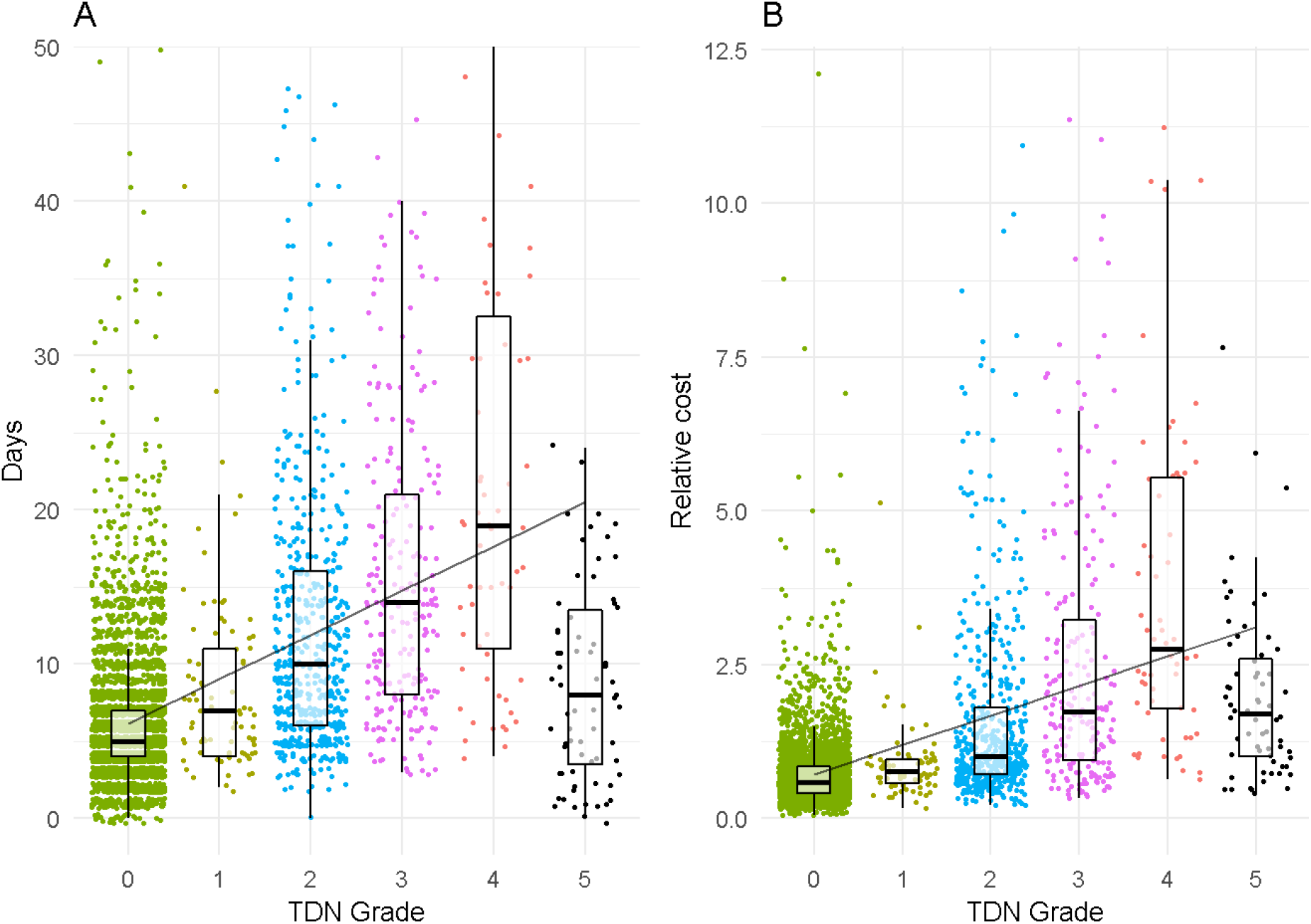
Length of hospital stay and relative cost at the University Hospital Zurich in relation to the Therapy-Disability-Neurology grade. Therapy-Disability-Neurology (TDN) grade = 0 means that no adverse event occurred prior to discharge. A. The y axis has been limited to values between zero and 50 days (20 values not shown). All values were included to compute the box plots. B. Relative cost corresponds to the ratio between the total cost of the case and the mean total cost of all cases.

At USZ, discharge and follow-up median KPS were lower in the group with AE (discharge: 70% [IQR 50]; follow-up: 80% [32·5]) than in the group without AE (discharge: 90% [10], median difference = 10% [10 – 20], p < 0·0001; follow-up: 90% [10], median difference = 10% [10 - 10], p < 0·0001). KPS at discharge was significantly affected by the TDN grade (p < 0·0001, Supplementary Figure 2). To analyze the outcome as a variation in functional status, we considered three categories: improved, unchanged, and worsened KPS at follow-up as compared to admission. The TDN grade significantly influenced the rate of each group (p < 0·0001) and a post hoc analysis of the mean ranks revealed that the outcome was worse as the severity of AE increased (Figure 5). This difference was significant for 7 out of 10 possible comparisons for TDN and 4 out of 10 for CDG (Supplementary Table 3). As admission KPS correlated with discharge KPS (tau = 0·56, p < 0·0001) and follow-up KPS (tau = 0·35, p < 0·0001), it could confound a correlation between absolute KPS values and AE grades. We therefore investigated the partial correlation between post-operative KPS and TDN grade by controlling for initial KPS and excluding grade 5 AE (death; always KPS = 0%), and obtained a significant negative correlation at discharge and follow-up (discharge: tau = - 0·23, p < 0·0001; follow-up: tau = - 0·22, p < 0·0001).

**Figure 5.**
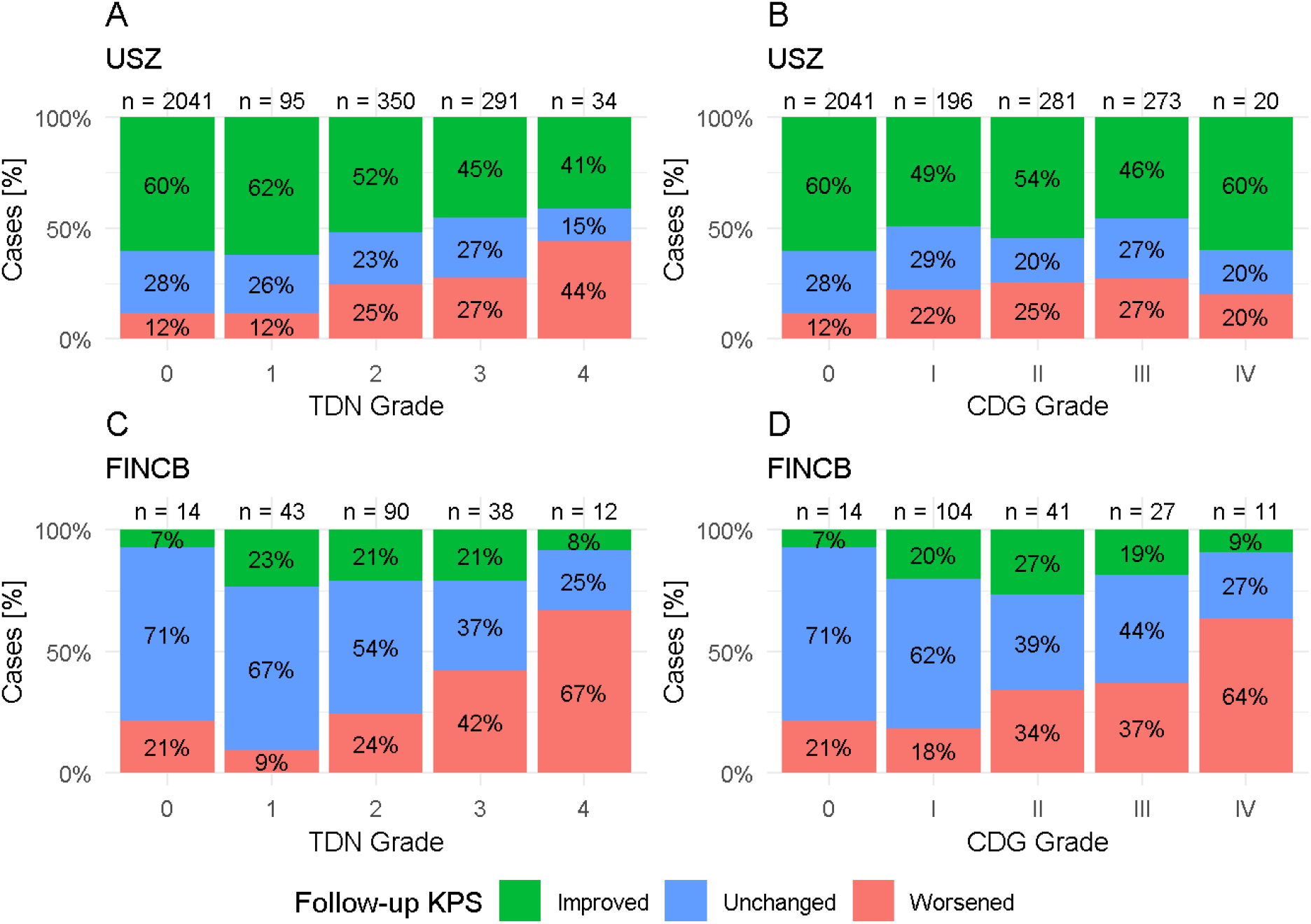
Karnofsky Performance Status Scale variation between follow-up and admission. The outcome was ordered into three groups: Karnofsky Performance Status Scale (KPS) improved (green), KPS unchanged (blue), KPS worsened (red) at follow-up as compared to admission. Grade 5 adverse events are always associated with a worsening of the KPS (data not shown). Grade 0 means that no adverse event occurred prior to follow-up. At University Hospital Zurich (USZ), follow-up consultations were recorded independently of surgical indication, for elective and urgent operations. At Fondazione IRCCS Istituto Neurologico Carlo Besta (FINCB), follow-up consultations were recorded only for patients who had an elective craniotomy. The KPS was more likely to remain unchanged at FINCB, due in part to the difference between the cohorts, and the fact that the admission KPS was higher at FINCB (Table 2). In both cohorts, each Therapy-Disability-Neurology (TDN) grade is associated with more cases with KPS deterioration and less cases with KPS improvement. This relationship is less consistent for the Clavien-Dindo-Grading system (CDG, for example, the outcome is worse for CDG I than for CDG IV in the USZ cohort). Statistical significance of the difference in the distribution of KPS variation between each grade of adverse event is given in Supplementary Table 3. Number of cases (n).

Multiple regression models using each dimension of TDN as predictors for LOS, costs and KPS at discharge (excluding grade 5 AE) showed that the dimension with the largest relative importance was “therapy” for LOS and relative cost, and “therapy” as wells as “disability” for discharge KPS (LOS: relative importance: therapy = 82%, disability = 13%, neurology = 5%, multiple R^2^ = 24%, p < 0·0001; relative costs: relative importance: therapy = 78%, disability = 17%, neurology = 6%, multiple R^2^ = 27%, p < 0·0001; KPS at discharge: relative importance: therapy = 49%, disability = 46%, neurology = 5%, multiple R^2^ = 17%, p < 0·0001). Linear regression models showed that TDN was as effective a predictor as the separate dimensions (LOS: R^2^ = 23%, p < 0·0001; relative costs: R^2^ = 25%, p < 0·0001; KPS at discharge: R^2^ = 13%, p < 0·0001).

Finally, we validated TDN against a dataset that was recorded independently at FINCB. The distributions of TDN dimensions differed between USZ and FINCB (Supplementary Figure 3). The correlation between LOS and TDN at discharge was moderate (rho = 0·53, R_s_^2^ = 28%, p < 0·0001, Supplementary Figure 4) and the partial correlation between absolute KPS values (at discharge and follow-up) and TDN, controlling for initial KPS and excluding grade 5 AE was moderate (discharge: tau = - 0·41, p < 0·0001; follow-up: tau = - 0·20, p < 0·0001). Using data from FINCB as a validation set for the linear regression models trained on data from USZ and described above, we found that variance in TDN could explain as much as 32% of variance in LOS and 27% of variance in KPS at discharge (LOS: R^2^ = 32%, mean absolute error (MAE) = 0·49; KPS at discharge: R^2^ = 27%, MAE = 12·85). Correlations obtained from FINCB external validation dataset were thus more pronounced than correlations obtained from USZ, and linear regression models were of comparable usefulness in both cohorts.

## DISCUSSION

Our results confirmed that AE significantly lengthen hospitalizations, increase costs and, most importantly, jeopardize short- and middle-term outcome of surgical neurology. At USZ we observed a moderate positive correlation between TDN and LOS as well as treatment cost. Each TDN grade was associated with a poorer outcome, independent of initial functional status. We validated the applicability of our classification using the same methods on an external dataset from FINCB and obtained even more pronounced correlations between TDN and LOS, as well as discharge/follow-up KPS. The TDN grade was thus a consistent measure of the severity of AE in two independent cohorts from two different countries, despite a different distribution of its dimensions (Supplementary Figure 3) and of the types of surgery (elective and urgent at USZ, elective only at FINCB).

A standardized identification, documentation, and classification of AE is a prerequisite to compare data across the literature and,^22,23^ despite several attempts at harmonization, their report remains highly inconsistent.^6,7,19,24-27^ In 2001, Bonsanto et al.^24^ classified AE according to their nature. Houkin et al.^7^, in 2009, categorized AE based on their nature, their cause and the morbidity and mortality in which they resulted. They subsequently divided them depending on their relation to the procedure, their predictability, and their avoidability. The adaptation of the CDG for neurosurgery, the LIC, was proposed in 2011.^6^ In 2016, Rampersaud et al.^25^ elaborated the Spinal Adverse Events Severity System version 2 (SAVES-V2). Their system assessed the severity of AE based on the therapy used to treat them, the duration of their consequences and the occurrence of severe neural injury. Albeit innovative, their integration of the consequences of AE was solely based on a subjective assessment of their likely duration and it did not account for the severity of the resulting disability. Castle-Kirszbaum et al.^26^ later modified the SAVES-V2 to include cranial events, but little change was made to the severity system. More recently, Gozal et al.^27^ classified AE in five categories based on their causes. While all these classifications were important contributions to the field, we believe that only a multidimensional system based on both the therapeutic and functional consequences of AE can adequately reflect clinical reality. Unfortunately, despite all efforts described above, no consensus was reached.

As a therapy-based classification scheme, the CDG has distinctive qualities that motivated us to integrate it in our patient registry, and in our new TDN grade. It has been broadly validated and it can be used as a basis to estimate the morbidity and predict the costs associated with multiple AE.^3-5,28,29^ The assessment of the severity of an AE based on the resources that have been deployed to counteract it is objective, and not influenced by the patients’ ultimate functional state. Consider a hardware malposition after the implantation of a neurostimulator. A re-operation that would increase the length of hospitalization would be necessary, but the patient may be discharged in great health. The CDG excels at capturing the magnitude of AE in such circumstances. A classification based solely on the disability of the patient at discharge would overlook the physical and moral harm, as well as the time loss and the cost generated by this event. In other instances - especially common in surgical neurology - the quality of a patient’s life can be seriously affected by an AE, without the need for treatment. For example, typical AE following surgery involving highly eloquent regions are focal neurological deficits.^10^ They can be highly disabling and it is in no way conceivable to consider such events as minor only because no treatment can be provided. The CDG ignores this subtility and has been shown to diverge from the NIHSS and KPS, especially for grade 1 AE (which includes most new neurological deficits).^8,10,14^ Therapy-only based classifications are robust options to assess the severity of AE, but lack the finesse needed to thoroughly capture the specifics of the neurological environment; these should be kept as pillars to support a more comprehensive grading scheme.

With the TDN grade, we complement the CDG formula by adding measures of functional disability and neurologic deficits. The widely used mRS,^30^ which quantifies disability effectively, is in its structure very similar to the CDG and constitutes its ideal complement. In a discipline in which the nervous system is of utmost importance, it could have been expected from our classification to integrate an extended classification of neurological functions such as the NIHSS, but we believe that functional independence is anatomically bound to the nervous system and that it represents an accurate reflection of its impairment. We chose not to duplicate these modalities to avoid redundancy and preserve the functionality and clarity of our classification. Still, we included a binary definition of neurologic deficits, as some conditions - for example sensory disorders - may decrease quality of life without impairing activities of daily living. Each of the stated dimensions of AE influences the quality of life of patients in a different way. The benefit of the TDN classification is that it combines them in a single numerical scale that correlates with both the clinical and economic repercussions of AE. Thus, the TDN grade retains the strengths of previous classification systems while providing greater accuracy and clinical adequacy.

The intent of the proposed classification is to determine the severity of AE and not to define them. For the purpose of this study, AE were defined as suggested by Dindo et al.^4^ Multiple other definitions have been proposed over the years, but they differ mainly in their ability to capture events with little impact on the post-operative course.^6,7,25^ We encourage a proactive discussion to reach agreement on this subject, and trust that our tool is sufficiently flexible to capture the severity of AE regardless of how they are defined.

Ideally, comparison of the occurrence and distribution of AE between different groups should be accompanied by measurement of surgical complexity. We strongly support the development and implementation of standardized scales to measure surgical complexity such as the Milano Complexity Scale (MCS).^19^ Nevertheless, the MCS is focused on brain tumor surgery and we were unable to apply it in this study, which covers a much broader spectrum of indications.

Classifying AE with the precision needed for surgical neurology is challenging; currently available tools minimize some highly invalidating conditions. As a solution, we propose an accurate, objective, and functional classification of the severity of AE that we evaluated with two of the largest data sets in the field. We hope that the Therapy-Disability-Neurology classification will be part of a shift towards a more standardized, critical, and patient-centered appraisal of adverse events, in an effort to provide safer surgical neurology.

## Data Availability

The data will be provided upon reasonable request.

## CONTRIBUTORS

MCN and JS formulated the study question. APRT, JS, and MCN designed the study and developed the new grading system. JS, MCN, and LR designed and maintained the Zurich neurosurgery patient registry. CMZ, SV, JV, FV, KA, SS, OB, PF, LR, MB, JS, and MCN collected data. APRT prepared the data. APRT, JS, and MCN did all statistical analysis. UH contributed to the study design and reviewed statistical analysis. All authors contributed to writing, data interpretation, critical revision of the work, and approved the final version.

## DATA SHARING

The data will be provided upon reasonable request.

## ACKNOWLEDGMENTS

We thank all physicians from our department as well as the administrative staff for rigorously including and reviewing patient data in our institutional registry.

## ABBREVIATIONS

AE: adverse event
CDG: Clavien-Dindo-Grading
CI: confidence interval
FINCB: Fondazione IRCCS Istituto Neurologico Carlo Besta
IQR: interquartile range
KPS: Karnofsky Performance Status Scale
LIC: Landriel Ibañez Classification
LOS: length of hospital stay
MAE: mean absolute error
MCS: Milano complexity scale
mRS: modified Rankin Scale
NIHSS: National Institutes of Health Stroke Scale
SAVES-V2: Spinal Adverse Events Severity System version 2
SD: standard deviation
TDN: Therapy-Disability-Neurology
USZ: University Hospital Zurich

## RESEARCH IN CONTEXT

### Evidence before this study

We searched PubMed for studies published in English until September 01, 2019, using the Title/Abstract terms “neurosurgery” OR “spine surgery”, AND “complication*” OR “adverse event*”, AND “classification” OR “classified” OR “graded”. Several classifications of neurosurgical adverse events have been proposed, but none have been widely reported in the literature. Adverse events were often recorded without a grade of severity or divided into minor and major according to various criteria. The most commonly used standardized schemes were the Clavien-Dindo-Grading system and the Landriel Ibañez Classification.

### Added value of this study

We present a novel patient-centered multidimensional classification of the severity of adverse events, along with a large two-center validation study on post-operative neurosurgical adverse events. Our results display the functionality of our approach through its correlation with length of hospitalization, treatment cost, and most importantly, patient outcome.

### Implications of all the available evidence

There is a high variability in the description of post-operative AE in surgical neurology. Standardization of this process would allow for a better comparison of the data reported and, as our study validates, is achievable in a consistent and functional manner.

